# Pathogenicity Assessment of a Bicuspid Aortic Valve–Associated *ELASTIN* Variant Using a Zebrafish Model

**DOI:** 10.64898/2026.05.05.26350845

**Authors:** Victor G. Bernard, Alexis Theron, Aurélien Drouard, Jean-François Avierinos, Chris Jopling, Stéphane Zaffran, Adèle Faucherre

## Abstract

Bicuspid aortic valve (BAV) is one of the most common congenital heart defects but its genetic basis remains incompletely defined. Extracellular matrix components play key roles in outflow tract (OFT) and valve development, but their contribution to BAV is not fully established. Following the analysis of a cohort of BAV patients, we identified a family harbouring a rare human *ELASTIN* (*ELN)* variant (p.Gln691X). To assess its pathogenicity, we generated a zebrafish *elna/b* double knockout (KO) using an RNAless CRISPR Cas9 strategy to avoid genetic compensation. This mutant exhibited cardiovascular defects including OFT anomalies, reduced stroke volume and dysmorphic aortic valves, highlighting Elastin’s critical role in cardiac development. We then used this model to test the *ELN* variant identified in the BAV family. We found that wild-type *ELN* mRNA was able to restore normal cardiac function and morphology, whereas the variant *ELN* mRNA failed to do so. This study establishes a robust *in vivo* model to assess *ELN* variant pathogenicity and provides evidence linking ELASTIN to BAV, opening new avenues for uncovering the genetic mechanisms underlying BAV.

## Introduction

Bicuspid Aortic Valve (BAV) is one of the most common Congenital Heart Defects (CHD), affecting 1–2% of the population and characterized by the presence of two instead of three aortic valve leaflets(Siu and Silversides, 2010). This malformation is often accompanied by secondary complications such as aortic stenosis, valvular regurgitation and aortic aneurysms, making it a significant public health concern (Hinton, 2012; Padang et al., 2012). Despite its high prevalence, the genetic landscape underlying BAV remains only partially understood, underlining the importance of identifying new molecular determinants to characterize its pathogenesis. The semilunar valve leaflets, including those of the aortic valve, develop from the remodelling of the outflow tract (OFT) cushions. As circulatory hemodynamic forces increase during cardiogenesis, smooth muscle cells adjacent to the OFT endothelium begin producing extracellular matrix (ECM) components that will strengthen and provide elasticity to the developing OFT and valve leaflets. Indeed, recent studies have highlighted the crucial contribution of ECM components to cardiac morphogenesis and homeostasis, providing not only structural integrity but also molecular signals that regulate endothelial and interstitial cell interactions during valve morphogenesis (Hinton and Yutzey, 2011). Consequently, in humans, disruption of ECM components has been linked to a variety of outflow tract pathologies such as aortic stenosis and BAV (Clift et al., 2022). One such ECM component is ELASTIN (ELN), which will provide elasticity and resilience to both OFT and valve leaflets (Schenke-Layland et al., 2004).

Pathogenic *ELASTIN* variants as well as reduced expression have been linked to supravalvular aortic stenosis (SVAS), a condition which leads to the narrowing of the aorta adjacent to the aortic valve (Merla et al., 2012). Although pathogenic *ELN* variants have not been directly linked to BAV, some studies have reported that patients with *cutis laxa* syndrome, caused by pathogenic autosomal-dominant variants in *ELN*, can also be affected by BAV (Hadj-Rabia et al., 2013; Lin et al., 2022). Furthermore, a chromosomal micro deletion which includes *ELASTIN* causes Williams Beuren syndrome which manifests with a variety of developmental defects including SVAS and BAV (Padang et al., 2012). Similarly, *Elastin* haploinsufficiency in mice also leads to progressive aortic valve degeneration (Hinton et al., 2010). Developmental studies indicate that in chick embryos, ELASTIN production is initiated at incubation day 3 (Hamburger-Hamilton stages 21) by cells that surround the endothelium of the aorta directly adjacent to the myocardium, before spreading throughout the vasculature (Rosenquist and Beall, 1990). In zebrafish, Elastin production in the OFT also commences at around 3 days post fertilisation (dpf) and coincides with cardiogenic events such as coordinated contraction and AV canal development that place increasing hemodynamic loads on the OFT (Faucherre et al., 2020).

Following a genetic screen of a cohort of BAV patients (Théron et al., 2022), we identified a family carrying a rare *ELN* variant, *ELN* (p.Gln691X). To test the pathogenicity of this variant, we developed a zebrafish *eln* knockout model. The zebrafish has emerged as a valuable model for studying cardiac development and CHD. Its suitability is reinforced by the routine use of CRISPR/Cas9-mediated genome editing to generate whole-organism genetic modifications (Tu and Chi, 2012). In addition, the optical transparency of zebrafish larvae during early developmental stages facilitates direct *in vivo* cardiac imaging. Notably, up to 82% of human disease-related genes are conserved in zebrafish, further supporting its translational relevance (Howe et al., 2013). Finally, previous studies have successfully employed zebrafish for pathogenicity testing, highlighting the robustness of this model for functional genetic analysis (Moreau et al., 2021; Odelin et al., 2023; Janin et al., 2023).

Here, we demonstrate that the zebrafish *eln* KO model we developed can be used to determine *ELN* variant pathogenicity. Using this approach, we show that the *ELN* (p.Gln691X) variant is likely pathogenic, providing a previously unreported link between *ELN* and BAV.

## Methods

This study was conducted in accordance with the principles of the Declaration of Helsinki. Patients were recruited between 2014 and 2018 in La Timone Hospital, Marseille, France. Patients were not included if they had connective tissue disease, previous aortic valve surgery and less than 18 years old. This study was performed in accordance with institutional guidelines. Patient recruitment and the entire study were approved by the “Comité de Protection des Patients Sud Méditerranée V, France (registration number: 2013-A01020-45). Written consent was obtained from all patients.

### Whole-exome sequencing (WES)

Peripheral blood samples were collected after obtaining written informed consent from all participating family members or their legal guardians. Genomic DNA was extracted using standard procedures. Family-based whole-exome sequencing was performed using the NimbleGen SeqCap EZ MedExome kit according to the manufacturer’s instructions (Roche Sequencing Solutions, Madison, WI, USA). Paired-end 150-bp reads were generated on an Illumina NextSeq 500 platform (Illumina, San Diego, CA, USA). Raw FASTQ files were aligned to the human reference genome (hg19; University of California Santa Cruz, UCSC) using the Burrows–Wheeler Aligner (BWA) with the Maximum Exact Matches algorithm. Alignment quality was assessed using Qualimap version 2.2.1.

Variant calling was performed using the Genome Analysis Toolkit (GATK), and variant annotation was carried out using ANNOVAR, following best-practice guidelines. Variant annotation and exome data analysis were further conducted using the Variant Annotation and Filtering Tool (VarAFT), version 2.17 (http://varaft.eu) (Desvignes et al., 2018).

### Variant prioritization

Variant prioritization was performed using VarAFT. A pedigree-based analysis was conducted considering autosomal dominant, autosomal recessive, X-linked dominant, and X-linked recessive modes of inheritance. In addition, both the “common disease–rare variant” and the “common disease–common variant” hypotheses were evaluated. This strategy was guided by the frequency of bicuspid aortic valve (BAV) in the general population.

### Zebrafish husbandry

Zebrafish were maintained under standardized conditions (Aleström et al., 2020) and experiments were conducted in accordance with the European Communities council directive 2010/63.

### Morpholinos

Morpholino oligonucleotides (MOs) were obtained from Gene Tools (Philomath, OR, USA) and injected into one-cell stage embryos. The sequences of the injected MOs are the following:

*elna* MO : 5′-GTTTCAGACCTCCAGCCATTGGGTA-3’ -5ng

*elnb* MO : 5′-GCCATCCTGCTCTGTAATAACCCTC-3’ -5ng

### Crispants

Crispants were obtained by co-injecting four non-overlapping gRNAs targeting either *elna*, or *elnb* with Cas9 protein into one-cell stage embryos. All gRNAs were obtained from Integrated DNA Technologies (Leuven, Belgium) and RNP complexes were assembled as described previously (Kroll et al., 2021). The sequences of the gRNAs are the following:

*elna* target sequences:

**1**-5′-CCTGGTGTAGCCACGGGGAC-3’;**2**-5′-TCCAAGCCTGGTTATCCAAC-3’**; 3**-5′-AGGACCCAGTCCTTGTACAC-3’**; 4**-5′-AGATTCTACCCAATGGCTGG-3’

*elnb* target sequences:

**1-** 5′-GAGCAGAGGTACCTTCCTCC-3’; **2**-5′-GGTCTACCAGGAATTGGAGC-3’; **3**-5′-TCCAACACCAGCCCCTAATC-3’; **4**-5′-GTACCAAGCCCTCCACCACC-3’

### Crispr/Cas9 “RNA less”

All gRNAs were designed to cut upstream and downstream of the transcription start site with two gRNAs per gene to impede RNA production and avoid genetic compensation (Hoshijima et al., 2019). The *elna* and *elnb* gRNAs were co-injected with nls-Cas9 protein (Integrated DNA Technologies).

*elna* target sequences:

**1**-5’-AGAGCTGTTGTAAATACTC-3’

**2-** 5’-GAGAAACATAACCAGCTTAT-3’

*elnb* target sequences:

**1**-5’-GAACGATTGAATGATGGGTC-3’

**2**-5’-AATGGCTTATTGTTGCAAGA-3’

### *elna/b* KO genotyping

Primers were obtained from Integrated DNA Technologies (Leuven, Belgium). DNA was extracted by lysing a zebrafish fin in 100µL of 50mM NaOH for 45 min at 95°C, followed by neutralization 10µL of 1M Tris-HCl. PCRs were performed using the PCR Master Mix from Promega (M7502)

*elna* primers:

Forward: 5’-CTGTTTTTACCCGACGCACT-3’

Reverse: 5’-TCCAAATTGAATGCCTCCTC-3’

*elnb* primers:

Forward: 5’-TTTGGGGCTAGAGGGGTTAT-3’

Reverse: 5’-TCATGTGAAGCCTCGAACAG-3’

### mRNA injection

The wild-type (WT) and mutant (p.Gln691X) Human *ELASTIN* cDNAs were subcloned into pCS2+ plasmids (GenScript). The plasmid was linearized using the *NotI* restriction enzyme and mRNA was synthesized using mMessage mMachine kit (Ambion). 100pg of mRNA was injected into single cell stage *elna/elnb* KO zebrafish embryos.

### Confocal imaging

The transgenic *Tg(myl7:EGFP), Tg(fli1ep:tdTomatoCAAX*) lines were crossbred to visualize outflow tract morphogenesis. Larvae were fixed at 5dpf using 4% paraformaldehyde and mounted in low-melting agarose (MilliporeSigma) on a glass coverslip to perform imaging with dual-channel confocal microscopy (Leica TCS-SP8-SMD, resolution 1024×512, 20X oil objective). Image acquisitions were performed at the MRI Facility.

### Blood flow analysis

To analyse zebrafish blood flow, we used the μZebraLab™ software (ViewPoint), developed for quantitative cardiovascular measurements. Experiments were performed as previously described (Faucherre et al., 2020). To determine the mean blood flow, the average of the blood flow values was calculated from a 10-second time frame (1,300 frames). Stroke volume was calculated by dividing the average blood flow (nL/sec) by the heart rate (BPM/60).

### Valve imaging

One day prior to imaging, larvae were incubated in 0.2 μM BODIPY-FL Ceramide (InvitrogenD3521) in Embryo medium + PTU (0.003% 1-phenyl-2-thiourea). 7dpf larvae were then anesthetized with Tricaine (0.16g/L) and mounted in low-melting agarose (MilliporeSigma) on a glass coverslip. Imaging was performed using a Zeiss LSM710 two-photon microscope coupled to a Ti:sapphire laser (Spectra-Physics, Santa Clara, CA, USA) and a water immersion 25× objective (Faucherre et al., 2020). Image acquisitions were performed at the IPAM Facility.

## Results

### Defective OFT and aortic valve in *eln a/b* KO zebrafish

Zebrafish possess two *elastin* paralogs, *elastin a* (*elna*) and *elastin b* (*elnb*) (Faucherre et al., 2020). We generated the double knockout (*elna/b* KO), using an “RNAless” approach in order to avoid genetic compensation due to mRNA decay (Hoshijima et al., 2019). We designed two gRNAs for each gene targeting sequences upstream and downstream of the transcription start site. This CRISPR/Cas9 strategy deletes the promoter region and prevents transcript production, generating an “RNAless” knockout line lacking *elastin* mRNA (Figure 1A). We initially assessed whether this approach had successfully deleted the target region by PCR. Wild-type *elna* and *elnb* alleles produced PCR fragments of 1800 bp and 1496 bp, whereas KO alleles produced shorter fragments of 1137 bp and 732 bp, respectively (Figure 1B). Next, we verified the absence of *elastin* transcripts by performing *in situ* hybridization using *elna* and *elnb* probes. In accordance with previous reports (Faucherre et al., 2020), we observed that both *elna* and *elnb* are expressed in the developing zebrafish OFT. In contrast, *in situ* analysis of *elna* KO and *elnb* KO larvae showed the complete absence of mRNA expression for both paralogs (Figure 1C). Because *eln* is expressed in the zebrafish OFT, we examined the structural integrity of this region by generating transgenic *elna/b* KO expressing tdTomato in endothelial cell membranes (Tg(*fli1ep:tdTomatoCAXX*)) and GFP in cardiomyocytes (Tg(*myl7:EGFP*)). At 4 dpf, the OFT has developed into a pear-like structure directly adjacent to the myocardium. However, in *elna/b* KO transgenic larvae, this structure failed to develop normally and was noticeably thinner/stenosed with a significant reduction in its width (Figure 1D, E). Next, we assessed cardiovascular physiology of *elna/b* KO larvae by performing blood-flow analysis at 4 dpf. Using this approach, we observed a significant reduction in the stroke volume of *elna/b* KO larvae (Figure 1F). Because aortic valves will form in the OFT and that alterations in cardiac function can be associated with valve abnormalities, we next examined aortic valve morphology at 7dpf using two-photon microscopy. In WT larvae, the valve leaflets appeared thin and flexible (See Movie 1). In contrast, 75% of *elna/b* KO larvae exhibited dysmorphic aortic valves, indicating that Elastin is required not only for OFT formation but also for proper aortic valve development in zebrafish (Figure 1G, H and see Movie 2). To independently validate that the phenotypes observed in *elna/b* KO larvae were specific, we adopted two additional strategies. First, we used morpholinos targeting *elna* and *elnb* to inhibit mRNA translation and, in parallel, we employed a “Crispant” approach (Kroll et al., 2021), by injecting four non-overlapping gRNAs targeting *elna and elnb*. Using these approaches, we determined that 80% of morphants and 84% of crispants exhibited aortic valve dimorphism, confirming our observations with *elna/b* KO larvae and supporting the suitability of our model for pathogenicity testing of human *ELN* variants (Figure 1G, H).

**Figure 1.**
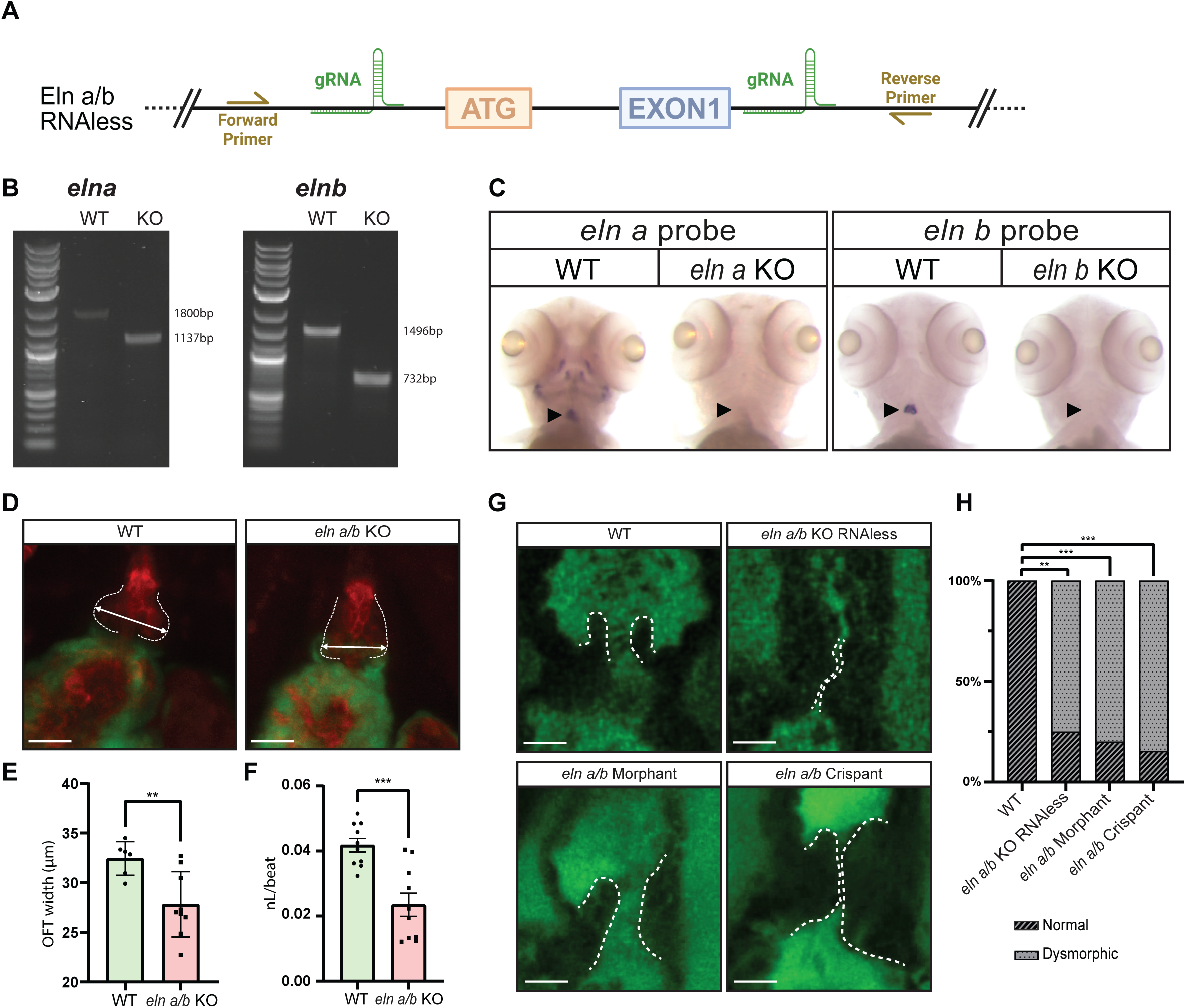
Inhibition of *elna/b* expression induces cardiovascular phenotypes. **(A)** Schematic representation of the gRNAs and primers localisation for generating and genotyping the RNAless *elna/b* KO line. **(B)** Agarose gel electrophoresis images showing genotyping of *elna* and *elnb* genes. Higher molecular weight bands correspond to WT, whereas the lower bands correspond to knock-out alleles. **(C)** Representative pictures of ISH against *elna* and *elnb*. Both *eln* expression can be detected in the OFT of WT but not of KO zebrafish larvae (black arrowheads) at 4dpf. **(D)** Confocal images of 4dpf Tg(*myl7:EGFP*), (*fli1a:tdTomatoCAAX*) larvae OFT from WT and *elna/b* KO. The white arrows indicate the width of the OFT immediately adjacent to the myocardium. Scale bar: 20 µm. **(E)** Graph associated with (D) showing the average size of the OFT of WT *elna/b* KO larvae (WT n=6, *elna/b* KO n=9. **(F)** Graph depicting the average stroke volume in nanolitres per heart beat (nL/beat) of WT (n=10) and *elna/b* KO (n=10). **(G)** Representative 2-photon images of the aortic valves of 7dpf zebrafish larvae labelled with BODIPY in WT, *elna/b* KO, *elna/b* crispants and *elna/b* morphants. Valves are outlined with a dashed white line. Scale bar: 15 µm. **(H)** Graph associated with (G) showing the quantification of aortic valve dysmorphism in WT (n=8), *elna/b* RNAless KOs (n=12), *elna/b* morphants (n=12) and *elna/b* crispants (n=13). ** p<0.01; *** p<0.001; Statistical values were obtained using the Student’s t-test (E, F) and Fisher’s exact test (H).

### *ELN* variant in a family affected by bicuspid aortic valve

By performing whole-exome sequencing, we identified an *ELN* variant in a BAV family with two affected members (Figure 2A). The index case (I-2) has a medical history notable for hypertension and underwent surgical aortic valve replacement with a 21 mm Magna Ease bioprosthesis for severe symptomatic aortic stenosis. Surgical observation confirmed a type 1 BAV with a heavy calcified raphe between the right and left coronary cusps (Sievers classification: Type I, R/L; international nomenclature: three sinuses, two leaflets, two commissures, and one raphe) (Michelena et al., 2021). No associated aortopathy was observed.

**Figure 2.**
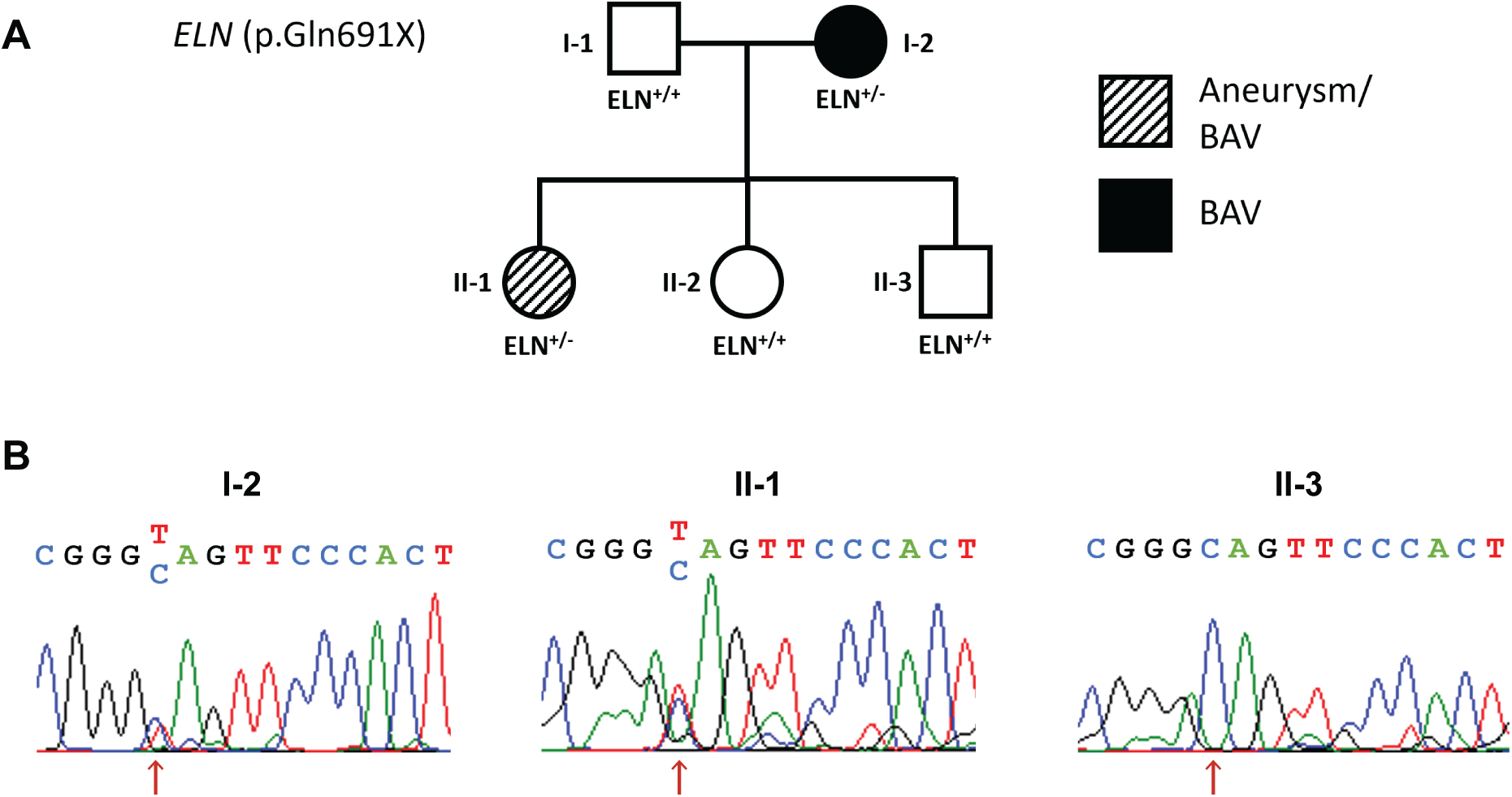
Family’s pedigree with *ELN* variant identification. **(A)** Family tree of patients suffering from BAV and carrying the *ELN* (p.Gln691X) variant. **(B)** Electropherogram of the mutation locus associated with three patients of this family. Genotypes are indicated below each symbol. (+) Reference allele; (-) Alternative allele.

Her daughter (II-1) has a history of treated hypothyroidism and was also diagnosed with aortic stenosis in the setting of a BAV with a raphe between the right and the left cusps, exhibiting a BAV phenotype identical to that of her mother. Aortic stenosis progressed to become symptomatic, with concomitant dilation of the ascending aorta that reached 41mm in diameter and displayed a tubular morphology. The patient subsequently underwent aortic valve replacement with a 19-mm Slimline Bicarbon mechanical prothesis, along with replacement of the tubular ascending aorta using a 26-mm Maquet Intergard vascular graft (Table 1).

**Table 1.** Clinical characteristics of patients with familial bicuspid aortic valve. Summary of clinical data of two related patients (mother I-2 and daughter II-1) with bicuspid aortic valve (BAV) and aortic stenosis. AVA, aortic valve area; LVEF, left ventricular ejection fraction; VKA, vitamin K antagonist.

| Characteristic | Patient I:1 | Patient II:1 |
| --- | --- | --- |
| **Sex** | Female | Female |
| **Relevant medical history** | Hypertension (treated with angiotensin II receptor blocker), chronic lymphocytic leukemia treated, stage 3 chronic kidney disease (vascular origin), Raynaud's syndrome, left facial paralysis | Treated hypothyroidism |
| **Aortic valve disease** | Severe symptomatic aortic stenosis | Progressive aortic stenosis monitored |
| **Bicuspid aortic valve phenotype** | Type 1 BAV with raphe between right and left cusps (Sievers I, R/L). Three sinuses, two leaflets, two commissures, one raphe | Identical phenotype: BAV with raphe between right and left cusps |
| **Pre-operative echocardiography** | AVA: 0.67 cm<sup>2</sup>; Mean gradient: 53 mmHg; LVEF: 65% | AVA: 0.84 cm<sup>2</sup>; Mean gradient: 44 mmHg; LVEF: 65% |
| **Associated aortopathy** | None | Ascending aorta dilatation (41 mm), tubular phenotype |
| **Cardiac surgery** | Surgical aortic valve replacement (in her 60's) | Aortic valve replacement and ascending aorta replacement (in her 50's) |
| **Valve prosthesis** | 21-mm Magna Ease bioprosthesis | 19-mm SLIMLINE BICARBON mechanical valve |
| **Aortic prosthesis** | None | 26-mm MAQUET INTERGARD tubular graft |
| **Follow-up (2025)** | Normally functioning aortic bioprosthesis, no structural valve deterioration, no aortopathy | No mechanical valve dysfunction, no prosthetic thrombosis, no hemorrhagic events under VKA therapy |

Whole-exome sequencing analysis allowed us to prioritize one stop-gain variant in the *ELN* gene (NM_000501:exon31:c.2071C>T; (p.Gln691X) ; MAF = 6.8 10^-6^ in the gnomAD database). This heterozygous rare variant was detected in the index case and her daughter. Subsequently, Sanger sequencing was performed for the family members and confirmed the segregation of the variant (Figure 2B). The variant is predicted to result in the loss of approximately 95 C-terminal amino acids, which contain hydrophobic domains conferring elasticity. *ELN*: (p.Gln691X) is predicted to be pathogenic by UMD-predictor(Salgado et al., 2016) and has a CADD-phred score = 44 (GRCh37-v1.6).

### Pathogenicity testing of the (p.Gln691X) human *ELN* variant

To determine whether the (p.Gln691X) *ELN* variant is pathogenic, we first assessed whether expression of human WT *ELN* mRNA could rescue the cardiovascular phenotypes we observed in *elna/b* KO zebrafish larvae. Stroke volume analysis shows that expressing human WT *ELN* mRNA in *elna/b* KO larvae restores this parameter to wild-type levels. In contrast, expression of the (p.Gln691X) human *ELN* variant mRNA failed to restore normal stroke volume, indicating that the *ELN* variant is unable to compensate for the loss of *elna/b* and is therefore potentially pathogenic (Figure 3A). We next assessed zebrafish aortic valve morphology at 7dpf. In this manner, we determined that expression of human WT *ELN* mRNA in *elna/b* KO larvae reduced the incidence of dysmorphic valves to 22%, demonstrating a substantial restoration of normal valve morphology. In contrast, expression of the (p.Gln691X) human *ELN* variant mRNA failed to rescue valve morphology (82% dysmorphic valves) (Figure 3B,C). Taken together, our data indicate that the (p.Gln691X) *ELN* variant is likely pathogenic and could therefore be associated with BAV in humans.

**Figure 3.**
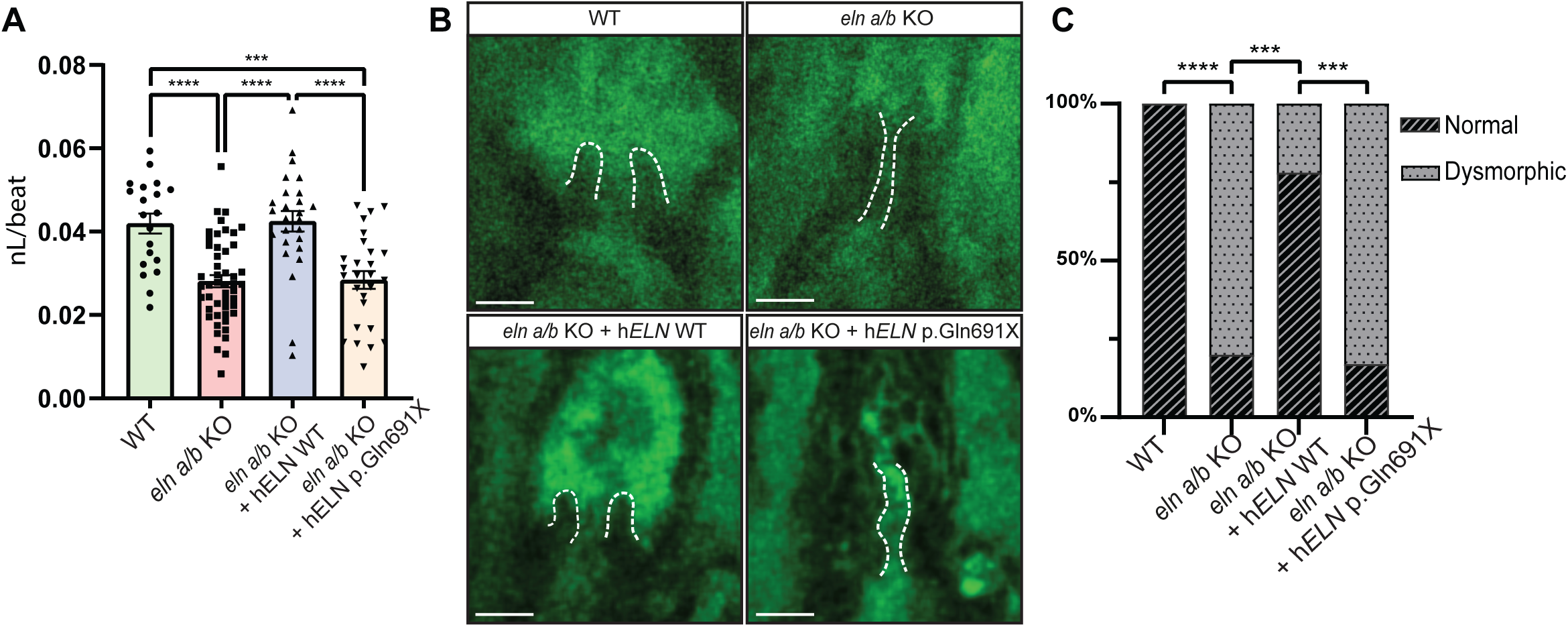
Pathogenicity testing of *ELN* mutant on zebrafish embryos. **(A)** Graph depicting the average stroke volume in nanolitres per heart beat (nL/beat) of WT (n=20) *elna/b* KO (n=49), *elna/b* KO injected with WT human *ELN* mRNA (n=26) and *elna/b* KO injected human *ELN* variant mRNA (n=27) **(B)** Representative 2-photon images of the aortic valves of 7dpf zebrafish larvae labelled with BODIPY in WT, *elna/b* KO, *elna/b* KO injected with human *ELN* WT mRNA and *elna/b* KO larvae injected with human *ELN* variant mRNA. Valves are outlined with a dashed white line. Scale bar: 15 µm. **(C)** Graph associated with (B) showing the quantification of aortic valve dysmorphism in a WT (n=10), *elna/b* KO (n=18), *elna/b* KO injected with human *ELN* WT mRNA (n=18) and *elna/b* KO larvae injected with human *ELN* variant mRNA (n=23). *** p<0.001; ****p<0,0001; Statistical values were obtained using one-way ANOVA followed by a Tukey’s multiple comparisons test (A) and Fisher’s exact test (C).

## Discussion

In this study, blood-flow measurements, OFT and aortic valve morphological analyses revealed marked cardiovascular defects in *elna/elnb* KO larvae. Dysmorphic valves were also observed in crispants and morphants, which not only validates our KO model but also enables other researchers in the field to adopt this approach to rapidly test variants’ pathogenicity without the need for acquiring and maintaining the *elna/elnb* KO line. Expression of human WT *ELN* mRNA consistently ameliorated the cardiovascular abnormalities observed in *elna/elnb* KO larvae, restoring functional and morphological parameters to levels comparable to WT larvae. In contrast, expression of human (p.Gln691X) *ELN* mRNA was unable to rescue these defects. Interestingly, while *ELN* mutations have classically been associated with SVAS, their involvement in BAV has not previously been described (Merla et al., 2012). Notably, the aortic valve abnormalities observed in *elna/b* KO larvae, characterized by thickened and dysmorphic valve leaflets, closely resemble those previously described when BAV-associated genes, including *notch1b* or *egfr*, are targeted in zebrafish larvae (Faucherre et al., 2020). This phenotypic concordance supports the hypothesis that *ELN* dysfunction may contribute to BAV pathogenesis. Notably, patients with *cutis laxa*, due to *ELN* defects, can develop BAV, further indicating that mutations in *ELASTIN*, either alone or in association with other mutated genes, may participate in broader molecular networks that contribute to BAV development (Hadj-Rabia et al., 2013). Beyond its role in valve development, *ELN* is a critical structural component of the aortic wall, where it confers elasticity, tensile strength, and resilience to cyclic hemodynamic stress. Elastin fibres are essential for proper lamellar organization of the aortic media and their disruption leads to progressive aortopathy. In humans, pathogenic *ELN* variants are classically associated with elastin-related arteriopathy, underscoring the sensitivity of the aorta to ELASTIN dosage and integrity (Merla et al., 2012). Our findings support a role for the *ELN* (p.Gln691X) variant in the BAV phenotype observed in the patients and suggest that it may also contribute to the concomitant abnormalities of the ascending aorta. Together, these results support a shared developmental and mechanobiological basis for both valvular and aortic disease in the context of *ELASTIN* deficiency.

Overall, our findings establish a robust *in vivo* framework that can be extended to the pathogenicity evaluation of other human *ELN* variants, including variants of uncertain significance (VUS), which would be of great interest for clinicians. However, while our findings provide functional evidence linking this *ELN* variant to valve defects, additional investigations will be required to definitively establish *ELN* as a causal gene in BAV.

## Data Availability

All data produced in the present work are contained in the manuscript

## Author contributions

AF and SZ were involved in study conception and design. All authors were involved in data collection and analysis. VB, AF, CJ and SZ drafted the manuscript. All authors contributed to the article and approved the submitted version.

## Acknowledgments

We thank the IPAM and MRI imaging facilities, members of the national infrastructure France-BioImaging (https://ror.org/01y7vt929) and supported by the French National Research Agency (ANR-10-INSB-04 & ANR-24-INBS-0005 FBI BIOGEN). CJ and AF are part of the Laboratory of Excellence Ion Channel Science and Therapeutics supported by a grant from the ANR. This work was supported by the “Fondation pour la Recherche Médicale” [DPC20111123002] and the “Institut National de la Santé et de la Recherche Médicale” to S.Z.

---

**Movie 1. Aortic valve in a 7dpf wildtype zebrafish larvae.**

2-photon imaging of a BODIPY labelled 7dpf zebrafish larvae reveals the valves dynamically moving during the cardiac cycle

**Movie 2. Aortic valve in a 7dpf *elna/b* KO larvae.**

2-photon imaging of a BODIPY labelled 7dpf zebrafish larvae reveals the valves dynamically moving during the cardiac cycle.

